# Maternal-Fetal immune networks and viral signatures in the healthy amniotic cavity

**DOI:** 10.64898/2026.06.17.26355783

**Authors:** M González-Rovira, L García-Díaz, C Martínez-Pancorbo, A Rodríguez-Herrera, J Sánchez, D Bernardo, M Karatas, JA García-Mejido, C Sousa, E Mellado, JA Sainz-Bueno, J Matthijnssens, ML Moreno

## Abstract

The intrauterine environment has traditionally been viewed as a privileged site protected by the placental barrier. However, emerging evidence suggests that early in utero microbial exposure may prime the developing fetal immune system. Here, using target-enriched metagenomics and high-dimensional proteomics, we characterized the intra-amniotic viral landscape and immune networks in 114 healthy pregnancies including both normal and anomalous fetuses. We identify a sparse yet heterogeneous human viral signature in 26% of samples, predominantly composed of *Herpesviridae*, *Polyomaviridae*, and *Picornaviridae*. Although viral reads abundance was associated with fetal abnormalities, viral detection generally did not induce overt inflammatory activation, supporting a state of immune homeostasis within the amniotic cavity. Instead, viral presence was associated with subtle and selective immune modulation, including altered inducible antimicrobial peptide expression (HBD-2 and HBD-3), coupled with an attenuation of regulatory cytokines. Our results further reveal that the amniotic immune environment is primarily governed by gestational age, transitioning from a Th1-predominant “alert” phase to innate-readiness preceding parturition. These findings suggest that fragments of viral genetic material within the amniotic cavity may contribute to fetal immune instruction without triggering overt inflammation, providing a foundational framework for understanding how “silent” viral-exposure during gestation influences the developmental origins of neonatal immunity.

## Introduction

The maternal-fetal interface serves as a critical barrier, integrating physical, molecular, and immunological mechanisms to safeguard the developing fetus (Yong et al., 2021). However, emerging evidence suggests that gestational immunity is not merely a maternal provision; rather, it involves a functionally competent fetal immune system capable of sensing environmental cues and contributing to the host defense response (Zhang et al., 2023). Contrary to the traditional view of fetal immune naivety, recent studies indicate that the fetus undergoes early in utero immune priming through exposure to microbial antigens, viral fragments, or maternal mediators that traverse the placenta (Mold et al., 2008; Arck and Hecher, 2013; Jennewein et al., 2017; Kollmann et al., 2017; Faas and Smink, 2025; Paarwater et al., 2025). Experimental models further demonstrate that fetal immune cells, particularly mononuclear phagocytes, can mount antigen-specific responses and actively participate in viral challenges (Abdelbasset et al., 2024).

Central to this environment, the amniotic fluid (AF) maintains intrauterine homeostasis through an intricate array of defence factors, including antimicrobial peptides (AMP), cytokines, immunomodulatory proteins, and vesicle-mediated signalling pathways (Kalbermatter et al., 2021; Kim et al., 2022; Bhatti et al., 2023). AMP such as HBD-1–3, HNPs 1–3, and LL-37, exert a dual role as both direct microbicidal effectors and potent immunomodulators capable of pattern recognition and fine-tuning inflammatory signalling (Šket et al., 2021; Fu et al., 2023).

While this complex network preserves homeostasis, protection is not absolute, and certain pathogens successfully evade these barriers (Lazzarotto et al., 2000; Mappa et al., 2023). The vertical transmission of pathogens such as cytomegalovirus (CMV), influenza, Zika virus and parvovirus B19 carries well-documented risks of malformation and severe complications, with outcomes largely dictated by gestational age at infection (Leruez-Ville et al., 2020; Megli and Coyre, 2022; Al Beloushi et al., 2024). In contrast, the detection of viral DNA or RNA in AF samples from asymptomatic or low-risk pregnancies remains infrequent, and the existing evidence is both scarce and heterogeneous, largely reflecting the absence of systematic routine screening (McLean et al., 1995; Baschat et al., 2003; Miller et al., 2009; Lim et al., 2018; Chen et al., 2024).

To date, research has primarily focused on the dysregulation of AMP and cytokines under pathological conditions, including preterm birth, intra-amniotic infection and inflammation, and preterm premature rupture of membranes (PPROM) (Tambor et al., 2012; Romero et al., 2015; Agakidou et al., 2022). Consequently, a notable knowledge gap persists regarding how these components maintain the pro- and anti-inflammatory balance during healthy gestations, particularly following the entry of viral fragments into the amniotic cavity. It is plausible that AF components actively neutralize viral particles or degrade free nucleic acids, resulting in transient, fragmented, or low-abundance viral signatures.

The investigation of these interactions in AF is constrained by the extremely low biomass of this compartment. Even prior to the advent of sequencing-based approaches, studies had demonstrated that microbial DNA detected in such environments frequently originates from environmental contamination or technical artifact rather than true biological presence (Salter et al., 2014; de Goffau et al., 2019; Olomu et al., 2020). Therefore, implementing rigorous controls ranging from stringent bioinformatic filtering to targeted validations is imperative to ensure that identified viral signals reflect genuine biological exposure rather than technical noise.

To address these gaps, the present study characterizes the AF ecosystem in clinically healthy pregnancies including both normal and anomalous fetuses by concurrently evaluating its viral composition and maternal-fetal immune networks. We sought to establish a baseline non-pathogenic viral profile, verify the biological validity of identified viral signatures, and investigate how AMP-mediated dynamics influence viral presence during normal gestation. Ultimately, elucidating the interplay between early viral exposure and the fetal immune response will optimize the interpretation of molecular diagnostics and provide novel insights into the developmental origins of immune function.

## Results

### Detection of *Herpesviridae* DNA in amniotic fluid samples

Although commercial viral immunoglobulin detection assays for human samples are currently available, their clinical utility depends on the immune activation caused by the virus and fails to detect the viral particle itself. To overcome these diagnostic limitations and given the well-documented association of these viruses with intra-amniotic inflammation and adverse pregnancy outcomes, we employed a highly sensitive PCR assay. Specifically, a total of 100 AF samples (n=85 from cohort 1, obtained by amniocentesis, and n=15 from cohort 2, collected during elective caesareans) from healthy pregnant women were screened for herpes simplex virus types 1 and 2 (HSV-1, HSV-2), CMV, and Epstein-Barr virus (EBV) via conventional PCR. This assay targeted the conserved DNA polymerase gene of this viral group using two sets of specifically designed primers. To ensure methodological rigor and exclude cross-contamination during amplification, negative controls were processed alongside each batch.

Among the primer sets evaluated, the H-G pair demonstrated the highest efficiency in amplifying the viral DNA polymerase gene. Among the 100 AF samples analyzed, screening revealed that six (6%) were PCR-positive, displaying heterogeneous amplification profiles (Fig. S1). Specifically, samples V11 and V74 exhibited prominent bands at approximately 500 bp, consistent with the expected molecular weights for HSV-1/2 and EBV. In contrast, samples V13, Q15, V63 and VR19 yielded larger amplicons (>500 bp), strongly points toward the presence of *Roseolovirus humanbeta6* (HHV-6A/B) and 7 (HHV-7) harboring hypervariable insertions that yield higher molecular weight PCR products.

Notably, four out of the six (66.7%) positive samples were associated with adverse fetal outcomes. Cases V11 and VR19 were linked to structural anomalies; specifically, V11 involved a diamniotic gestation complicated by preterm premature rupture of membranes (PPROM) in the presence of subclinical intra-amniotic infection. Furthermore, cases V74 and V13 were associated with chromosomal abnormalities. The remaining two PCR-positive cases presented no identifiable fetal pathologies.

Following the detection of *Herpesviridae* in 6% of the AF samples, we expanded the analysis to encompass a broader range of viral families through metagenomic sequencing, thereby increasing analytical depth and strengthening the reproducibility of the results. However, because conventional PCR amplification had depleted the residual sample volume, subsequent targeted molecular analyses could only be performed on sample V11 and on those study samples with sufficient remaining volume (Table S1).

### Metagenomic enrichment for the characterization of the amniotic fluid viral signature

To further characterize the viral landscape within the AF samples, we employed a metagenomic approach using a capture-based target enrichment protocol (Twist Bioscience). This strategy was specifically designed to enhance the signal-to-noise ratio of viral genetic material, thereby enabling the identification of a broad spectrum of viral species and providing a detailed map of viral diversity in the AF samples.

A subset of 50 AF samples was randomly selected for in-depth metagenomic characterization. Metagenomic analyses were performed on these samples (n=50) together with negative controls (n=7). To minimize cross-contamination and sequencing carry-over effects, stringent filtering criteria were applied, requiring a minimum of 100 sequencing reads, a Reads Per Million (RPM) value ≥ 1, and at least 500 covered bases for each detected taxon. This multi-step strategy was designed to rigorously distinguish genuine biological signals from technical artifacts, such as spurious mapping of reads, index hopping, background noise, and laboratory-derived cross-contamination. A first notable discrepancy in HSV-1 detection was observed when comparing RPM thresholds of 0.1% (ten samples) and 1% (samples V11 and VR48), thereby requiring additional technical validation. The qPCR confirmed positivity exclusively in samples V11 and VR48, demonstrating total concordance with the 1% filtering threshold. On the other hand, a high read counts of rotavirus were detected in the analyzed samples (27/50). Given that samples from another study with a very high rotavirus viral load were included in the same sequencing run, an independent qPCR validation targeting the NSP3 gene was performed, yielding negative results.

The implementation of stringent filtering criteria, based on the 1% RPM threshold, proved essential as a robust indicator of authentic viral presence, thereby establishing it as the standard for all downstream analyses. This bioinformatic pipeline resulted in the recovery of viral sequences in 16/50 AF samples (32%). These detections were distributed between second trimester (68.7%) and third trimester (31.3%) AF samples. Most reads were assigned to human-associated viral families, though plant viruses were also present. The viral landscape was characterized by a non-uniform distribution across sequencing reads, taxonomic abundance, and genomic coverage (Fig. 1). Specifically, the majority of taxa appeared at low abundance, with high-intensity viral signals, defined by elevated read counts and extensive coverage, limited to a small subset of samples.

**Fig. 1.**
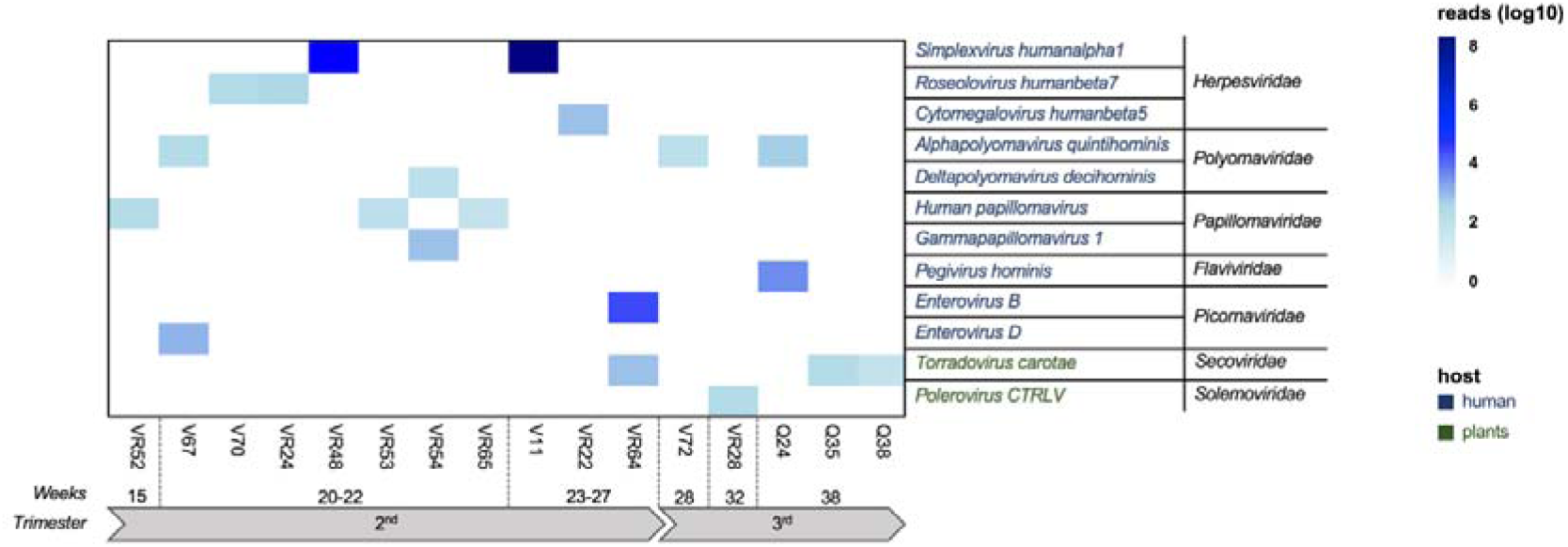
Viral landscape after filtering and species detection in amniotic fluid. Heatmap showing the distribution of viral species in the 16/50 amniotic fluid samples selected after filtering. Rows show viral species with their respective families, and columns indicate samples ordered by gestational age. Species nomenclature is color-coded according to primary host (human or plant).

We stratified the viral profiles detected in 16 positive samples out of the 50 analyzed by comparing them with documented intrauterine pathogens and clinical phenotypes. As illustrated in Figure 2, the human viral landscape comprised both, already known and novel signatures categorized by genomic structure. Regarding human-associated viruses, we observed a bifurcated distribution: enveloped DNA viruses (*Herpesviridae*) were predominantly associated with structural malformations, whereas non-enveloped DNA viruses (*Polyomaviridae* and *Papillomaviridae*) were identified in both healthy gestations and those with fetal pathology. Notably, *Picornaviridae* (RNA) signatures correlated with early-onset intrauterine growth restriction (IUGR), while members of the *Flaviviridae* were restricted to normal development. Among non-human viruses, the plant-associated taxa (*Secoviridae* and *Solemoviridae*) showed no specific clinical correlation, appearing in cases ranging from normal development to severe hydrops fetalis (Fig. S2). Subsequent comparative analyses between virus-positive and virus-negative AF samples were restricted to the 13 AF samples presenting human-associated viral signatures, as the scope of this study specifically focuses on the pathophysiological implications of these agents within clinically healthy pregnancies.

**Fig. 2.**
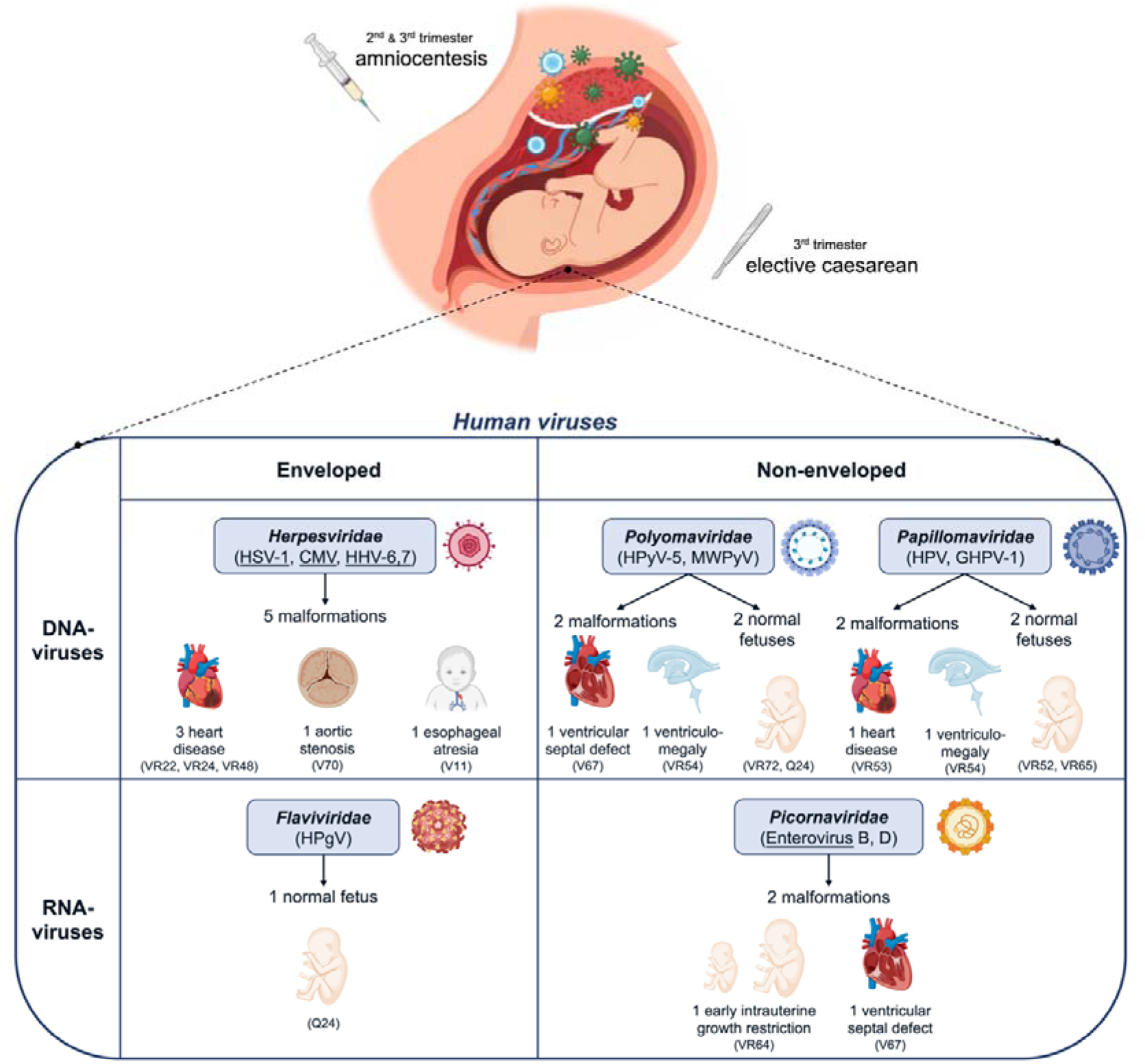
Classification of viruses detected in the intrauterine environment and associated clinical outcomes. Human viruses detected in 13 amniotic fluid samples that passed quality filtering among the 50 samples analyzed. Human viruses classified by genome type (DNA or RNA) and envelope status (enveloped vs non-enveloped), alongside their respective clinical outcomes. Underlined species denote viruses with previously demonstrated transplacental transmission capability. CMV, *Cytomegalovirus humanbeta5*; GHPV-1, *Gammapapillomavirus1*; HHV-6,7, *Roseolovirus humanbeta6,7*; HPgV, *Pegivirus hominis*; HPV, *Human papillomavirus;* HPyV-5, *Alphapolyomavirus quintihominis*; HSV-1, *Simplexvirus humanalpha1*; MWPyV, *Deltapolyomavirus decihominis*.

### Antimicrobial peptides concentrations in amniotic fluid samples

A total of 63 AF samples were selected from the initial pool of 108 recollected AF samples to provide representation across the different sample groups included in the study for the quantification of five specific AMP: HBD-1, HBD-2, HBD-3, HNPs 1-3, and LL-37. As illustrated in Fig. S3a, all targeted AMP were detectable across the study group, although marked interindividual variability was observed. HBD-1 and HBD-3 exhibited the highest overall concentrations, with values frequently reaching the 10^4^pg/ml range. In contrast, HBD-2 displayed a wider dispersion, characterized by numerous low-level measurements. While HNPs 1-3 and LL-37 showed lower overall medians, they exhibited sporadic high-value outliers, further underscoring the heterogeneous nature of AMP expression in the amniotic environment.

For subsequent comparative analysis, samples were stratified based on fetal clinical classification into normal development (n=31) and congenital anomalies (n=32), including malformations and chromosomopathies (Fig. 3a). Sample V11 (highlighted in red), a previously confirmed HSV-1 positive AF sample included as a subclinical infection control, was excluded from this analysis to avoid bias. HBD-1 levels were comparable between groups (*p*=0.5199), indicating constitutive expression independent of clinical status. Similarly, neither LL-37 (*p*=0.1319) nor HNPs 1-3 (*p*=0.2121) showed significant variation. In contrast, HBD-3 was significantly elevated in the anomalies group (*p*=0.0042), while HBD-2 also showed a significant but less marked increase (*p*=0.0498).

**Fig. 3.**
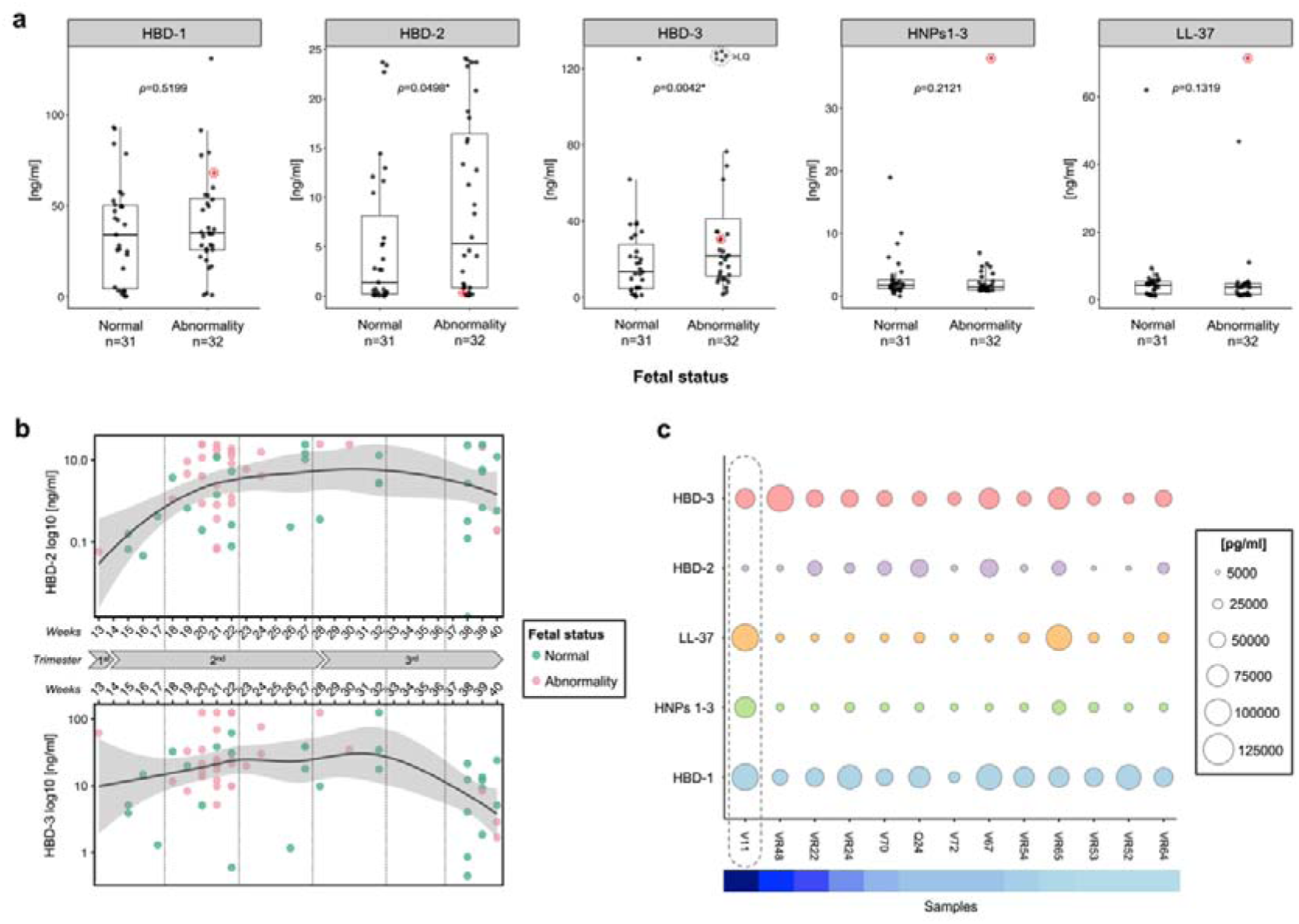
Antimicrobial peptide profiles across pregnancy and fetal conditions. Data correspond to 63 samples. **a** Comparison of the specific five AMP (HBD-1, HBD-2, HBD-3, HNPs 1-3, and LL-37) concentrations (pg/mL) stratified by fetal status (normal vs. abnormality). Boxplots with overlaid individual data points are shown for each peptide. Statistical significance was determined using the non-parametric Mann-Whitney U test; *p*-values are indicated above each comparison. Red circles highlight sample V11, a subclinical Herpesvirus-positive case. **b** Longitudinal dynamics of inducible defensins across gestational age ranges (13-17, 18-22, 23-27, 28-32 and 37-40). Smoothed curves represent the gestational trends of HBD-2 and HBD-3 concentrations on a logarithmic scale. Grey shading denotes the 95% confidence interval (CI), and vertical dashed lines indicate trimester boundaries. Green points represent AF samples from normal fetuses, while pink points represent AF samples from fetuses with abnormalities. **c** Bubble plot summarizing AMP abundance in the 13 samples with detectable human viral presence. Rows represent peptides and columns denote individual samples. Bubble size is proportional to peptide concentration (pg/mL), illustrating inter-individual variability and peptide-specific distribution patterns. The dashed line indicates the subclinical infection control sample. HBD, human beta-defensin; HNP, human neutrophil peptide.

Samples were next stratified by gestational age ranges (13-17, 18-22, 23-27, 28-32, and 37-40 weeks) to account for the rapid physiological changes occurring during fetal development. This stratification approach enhances statistical power within each subgroup and enables inter-group comparisons that more accurately reflect the distinct physiological stages of the second and third trimesters. Significant differences were observed exclusively for HBD-2 and HBD-3 (*p*=0.0114 and *p*=0.0021), suggesting a gestational influence specifically on these inducible defensins (Fig. 3b). This analysis showed that HBD-2 remained low in early gestation period, followed by a progressive increase starting around week 18 and stabilizing from week 27 onward. In contrast, HBD-3 peaked during mid-gestation and declined in late pregnancy, particularly in elective caesarean samples (37-40 weeks). Both peptides showed considerable variability across time points.

We next assessed the influence of fetal sex (male vs. female) and maternal age on amniotic AMP concentrations. Fetal sex showed no significant effect on most amniotic AMP, except for HNPs 1–3, which were higher in males (*p*=0.0063) (Fig. S3b). Maternal age (≥35 vs <35 years) had no significant impact on AMP levels, indicating that these immune markers are largely independent of maternal age in this cohort (Fig. S3c). Similarly, analysis of the AF samples with detectable viral presence (n=12) vs. no detectable virus (n=37) revealed no significant differences in AMP concentrations (Fig. S3d). V11 was also excluded from this analysis to avoid bias.

Sample-level visualization within the virus-positive subgroup confirmed marked inter-individual heterogeneity in AMP expression. HBD-1 was the most consistently abundant and stable peptide across the study group. In contrast, HBD-3 exhibited pronounced peaks in a subset of individuals, suggesting individual-specific high-level expression. While HBD-2 showed moderate variability, HNPs 1-3 and LL-37 displayed generally uniform distributions, albeit with occasional outliers (Fig. 3c). An antagonistic regulatory pattern was identified among inducible β-defensins, characterized by a divergence in HBD-2 and HBD-3 expression. Several cases exhibited an inverse expression profile, where increased HBD-3 levels were associated with HBD-2 depletion. This reciprocal dynamic was corroborated by Spearman correlation analysis, which revealed a weak negative association (ρ=−0.28). Although this trend did not reach the conventional significance threshold, the data suggests a mechanism of functional compensation or differential regulation orchestrated by exposure to viral genetic material. Additional sub-analyses accounting for genome type (DNA vs. RNA) and envelope status (enveloped vs. non-enveloped) yielded no significant results (Fig. S3e).High-throughput profiling of cytokines and inflammatory proteins in amniotic fluid using Olink proteomics

For the Olink proteomic analysis, we included a total of 62 AF samples, comprising the 50 samples subjected to metagenomic sequencing plus additional 12 random samples to ensure a comprehensive representation of the sample set (Table S1). Comparative analysis between trimesters revealed differential expressions of several cytokines (Fig. 4a). Specifically, markers associated with innate and cytotoxic responses (IL-1A, *p*=5.24e-11; IL-1B, *p*=0.0023; XIAP, *p*=0.0012) and leukocyte recruitment (CCL-20, *p*=0.0026) were significantly elevated in third-trimester samples. Conversely, second-trimester samples exhibited a predominance of Th1-related cytokines (IL-12A/IL-12B, *p*=4.34e-05; IL-12B, *p*=0.0002; TNF, *p*=0.0002) and Th2 mediators (IL-4, *p*=0.0013), alongside additional signatures of innate/cytotoxic activity (TGFB-1, *p*=0.003; GZMB, *p*=4.64e-04; GZMH, *p*=0.0386). No other immune mediators showed significant differences. Although sample V11 was removed from the statistical analysis to avoid bias, it was included in the figure to illustrate the distinction of these cytokines during a viral subclinical infection.

**Fig. 4.**
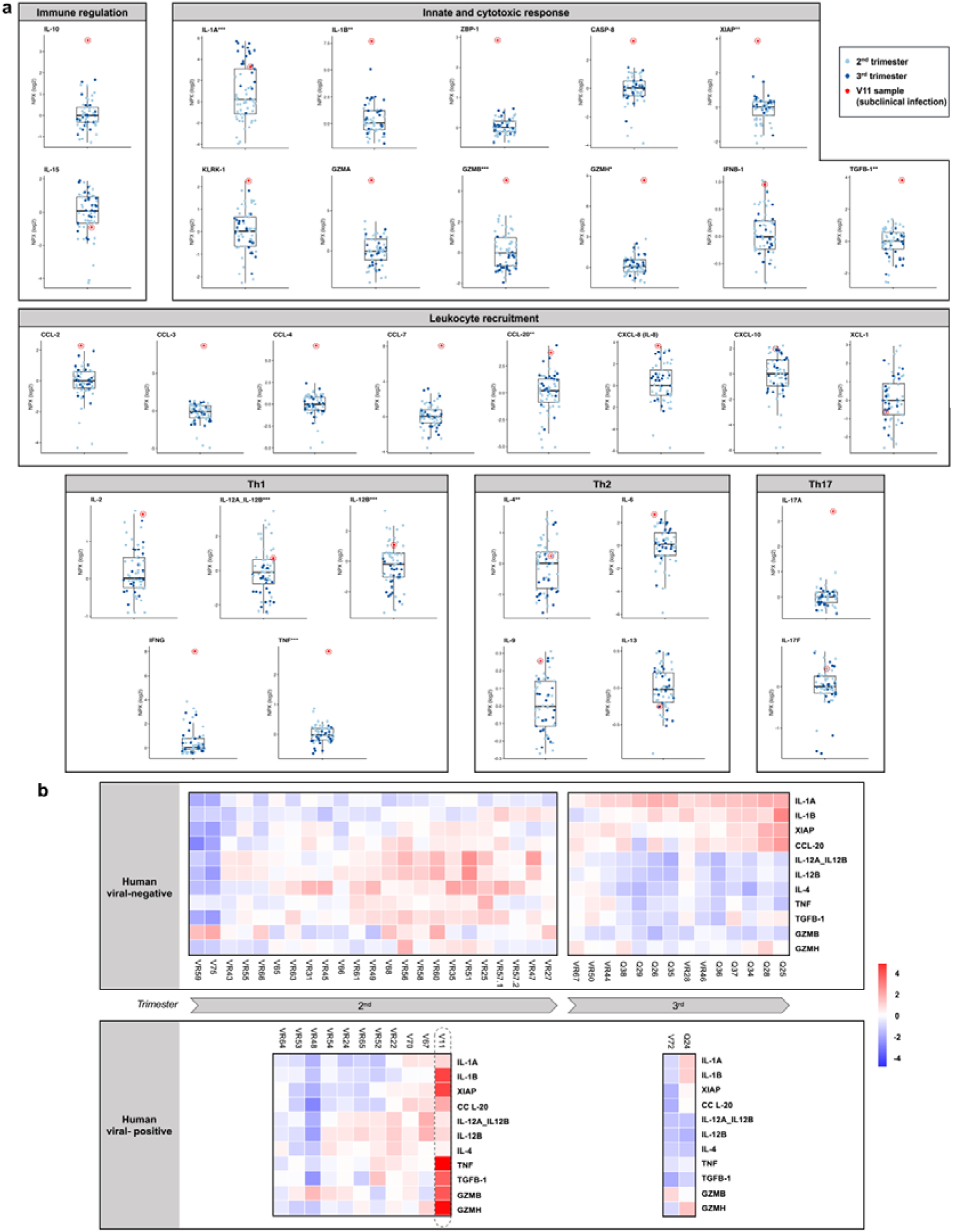
Immunome profile of the human amniotic fluid. 62 AF samples were included in the Olink proteomic analysis. **a** Immune mediators related with the innate and cytotoxic response, immune regulation, leukocyte recruitment and Th1, Th2 and Th17 immune profile were determined in the human amniotic fluid during the second (n=41) and third (n=21) trimester of pregnancy. Boxplots represent Normalized Protein Expression (NPX) values on a log2 scale, with individual data points overlaid (light blue: 2nd trimester; dark blue: 3rd trimester). Statistical significance between trimesters is indicated by asterisks (∗*p*<0.05, ∗∗*p*<0.01, ∗∗∗*p*<0.001). Red circles highlight sample V11, a subclinical Herpesvirus-positive case, as a representative outlier with the greatest deviation from the cohort distribution. **b** Heatmap displaying the eleven immune mediators with significant differential expression between trimesters. Samples are stratified by gestational age and viral detection status (human viral-negative vs. human viral-positive). The dashed line indicates the subclinical infection control sample.

Beyond gestational age, we evaluated the impact of other clinical variables on the amniotic inflammatory profile. While fetal sex showed no influence on cytokine levels, stratification by clinical status revealed significant alterations. Specifically, markers of leukocyte recruitment (CXCL-10, *p*=0.0325), Th1 response (IL-12A/IL-12B, *p*=0.0010; IL-12B, *p*=0.0006), and Th2 signalling (IL-4, *p*=0.0027) were significantly elevated in cases of fetal pathology (Fig. S4a). This upregulation followed a consistent pattern, with the highest concentrations identified in fetuses with structural malformations, followed by those with chromosomopathies, compared to normal-development controls. Conversely, women of advanced maternal age (≥35 years) showed increased pro-inflammatory signalling (IL-1B) during the second trimester (Fig. S4b).

To evaluate the relationship between proteomic profiles and viral presence, we restricted our analysis to the 50 samples with available metagenomic data. A heatmap of the eleven trimester-associated cytokines (Fig. 4b) confirmed that gestational trends remain robust, regardless of viral status. Interestingly, a subset of virus-positive second-trimester samples (V11, V67, and V70) exhibited elevated innate/cytotoxic activity (specifically IL-1A) at levels comparable to the third trimester. Notably, a distinct cluster (V75, VR59, VR48, VR53, and V72) displayed a lack of cytokine activation, except for the cytotoxic mediator GZMB. This was most striking in sample VR48, which failed to elicit a measurable inflammatory response despite harbouring a substantial viral load (∼20,000 *Simplexvirus* reads).

When comparing the immune landscape between groups, viral-positive samples paradoxically exhibited significantly lower levels of XCL-1 (*p*=0.0174) (Fig. S4c). These findings indicate an attenuation of regulatory and effector molecules in the presence of viral DNA, supporting the notion that viral detection in AF, excluding active infection cases like V11, may be associated with a state of immune quiescence or evasion rather than overt activation.

### A multi-platform analysis by integrating viral NGS/PCR and maternal-fetal proteomics

A multi-platform integrative analysis combining metagenomic viral detection, targeted *Herpesviridae* PCR validation, AMP profiling, and maternal-fetal proteomic characterization was subsequently performed across the 42 selected samples. Samples were organized according to viral detection status and gestational trimester in order to evaluate coordinated host-microbial signatures within the intra-amniotic environment. Viral read abundance varied substantially across positive samples and detected viral species. A small subset of viral signatures accounted for the highest log10-transformed read counts, whereas most detections were characterized by low-to-moderate abundance. These findings indicate that the amniotic viral landscape is highly heterogeneous, with discrete viral signatures rather than a generalized increase in viral reads across all positive samples.

Analysis of AMP revealed heterogeneous abundance patterns across the AF samples (Fig. 5b), with distributions remaining relatively stable throughout gestation. A fully coordinated anti-viral response is typified by the subclinical infection control sample V11, which serves as a positive benchmark where high read counts synchronize with the maximal, concurrent activation of both the innate chemical barrier (specifically LL-37, HNPs 1-3 and HBD-3) and a global pro-inflammatory cytokine storm spanning innate, chemotactic (IL-1B, CASP-8, CCL-7, CXCL-8), and T-helper (IFNG, TNF, IL-17A) pathways. However, localized variations were also evident; a distinct spike in LL-37 was observed in sample VR65. Intriguingly, the beta-defensins HBD-1 and HBD-3 were detected irrespective of viral load or gestational age. Consistent with its role as a constitutive baseline peptide largely independent of infectious or inflammatory cues, HBD-1 expression was broadly preserved across samples, with a prominent baseline elevation observed in sample VR59. Notably, sample V72 lacked detectable constitutive HBD-1 expression despite belonging to the viral-positive group, suggesting that viral detection was not uniformly associated with AMP activation. Conversely, elevated HBD-3 abundance appeared to cluster preferentially in samples associated with congenital malformations.

**Fig. 5.**
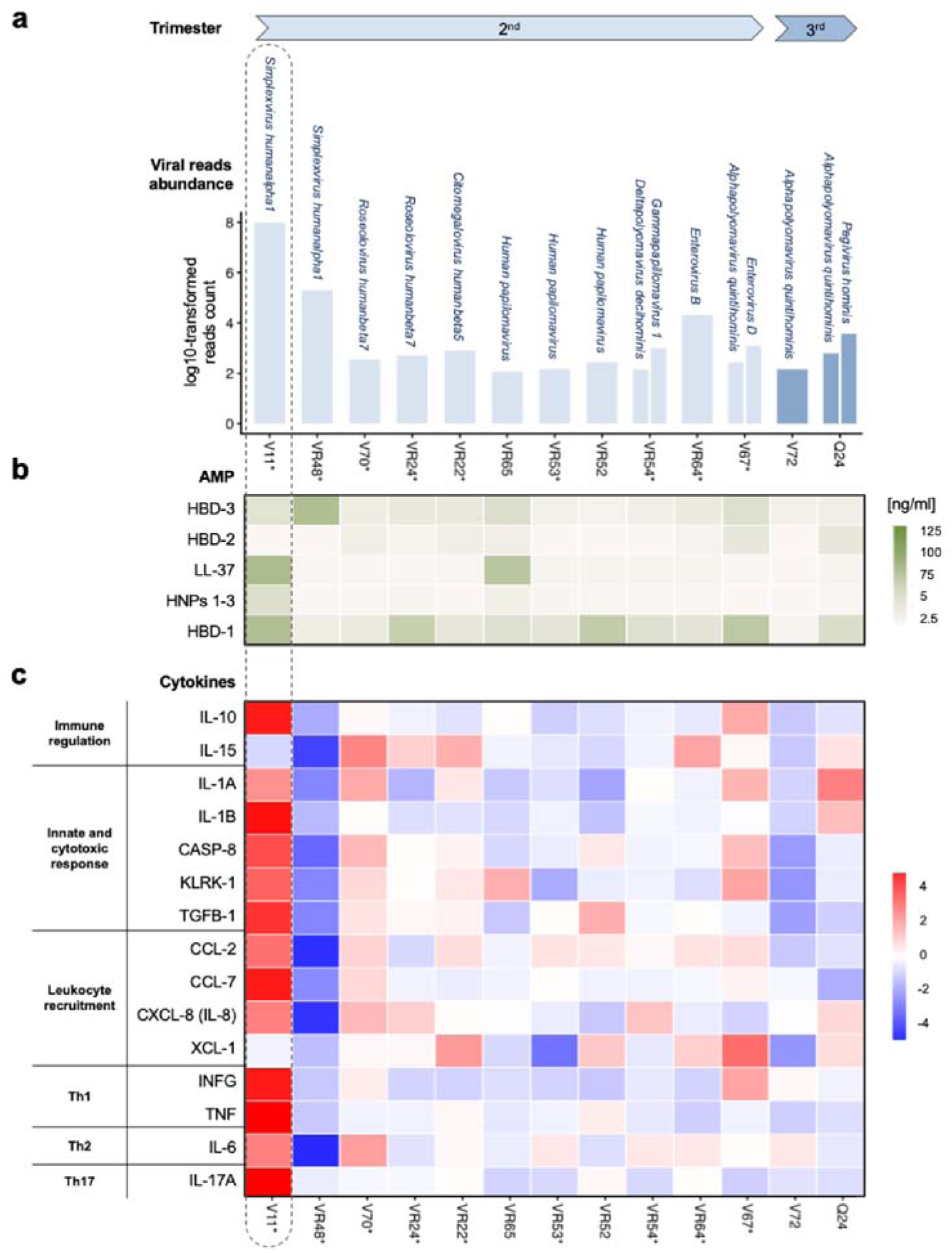
Multi-platform integration of viral detection and maternal-fetal immune profiling. **a** Viral reads abundance across the 42 selected samples, stratified by gestational trimester. **b** Heatmap of AMP abundance across trimester. **c** Heatmap of cytokine/proteomic markers across trimester. Asterisks denote samples from fetuses with anomalies (chromosomopathies + malformations). The dashed line indicates the subclinical infection control sample. AMP, antimicrobial peptides.

Maternal-fetal proteomic profiling further demonstrated the absence of overt inflammatory activation in viral-positive samples (Fig. 5c). Cytokine expression patterns were highly heterogeneous across individuals and did not segregate according to viral detection status. Classical inflammatory mediators, including IL-1B, CXCL-8, IFNG, TNF and IL-6 displayed variable expression across both viral-positive and viral-negative groups, without evidence of coordinated upregulation associated with viral detection. Notably, sample V72 showed no broad AMP and cytokine activation despite belonging to the viral-positive group, except for isolated IFNG elevation. Finally, VR48 displayed a lack of cytokine activation. These findings further support the absence of a generalized inflammatory response associated with the detection of viral genetic material within the amniotic compartment.

## Discussion

The presence and biological significance of viral nucleic acids within the AF remain poorly characterized, particularly in the context of healthy pregnancies. While understanding in utero microbial exposure is foundational to neonatal health (Mogotsi et al., 2024), unbiased metagenomic characterizations of AF viruses remain strikingly sparse (Baschat et al., 2003; Reddy et al., 2005; Lim et al., 2018). By integrating targeted PCR, enriched metagenomics, and high-dimensional immune profiling, our study provides a high-resolution map of the intra-amniotic viral landscape. Our findings reveal a sparse and heterogeneous viral signature, characterized by a notable absence of robust immune activation within the maternal-fetal environment. These data suggest that, while viral elements may transit into the amniotic compartment, the intrauterine environment maintains a state of immunological quiescence under homeostatic conditions.

Several mechanistic scenarios may account for the trace levels of viral genetic material observed within the amniotic compartment. First, these low-abundance signatures may capture the resolution phase of a subclinical, acute intrauterine infection, wherein residual or non-replicative genomic fragments persist despite the clearance of infectious virions (Gervasi et al., 2012; Chudnovets et al., 2020). Second, they could reflect sampling at an incipient stage of the viral lifecycle, preceding the exponential phase of replication within the amniotic cavity. Alternatively, rather than indicating an active intra-amniotic infection, these nucleic acids may represent vertical shedding from systemic or compartmentalized maternal reservoirs; such fragments could cross the maternal-fetal interface via hematogenous dissemination, cellular trafficking, or microvesicle-mediated transport without establishing a productive, high-titer fetal infection (Romero et al., 2015; Lee et al., 2019).

Specific taxonomic identification further underscores the complexity of this interface. Utilizing targeted PCR, we detected herpesvirus DNA in 6% of AF samples, a prevalence that marginally exceeds previous reports for obstetric cohorts (Romero et al., 2014; 2015). Despite methodological divergence between studies, our results confirm that herpesviruses can infiltrate the amniotic compartment during clinically asymptomatic pregnancies. This suggests that these viruses may possess specialized mechanisms to circumvent the robust physical and immunological checkpoints of the placenta (Beltrami et al., 2023), potentially through direct trophoblast infection or localized immune modulation (Parums, 2024; Salem et al., 2026).

Interestingly, the detection of viral nucleic acids, even in cases associated with adverse fetal outcomes, was not accompanied by a detectable inflammatory response in the AF. This dissociation between the presence of viral signatures and the activation of the immune response represents one of the most relevant findings of our study. Our results suggest that the intra-amniotic viral burden may remain below the threshold required to trigger fetal innate immunity or, alternatively, may reflect a specialized state of immune tolerance that protects the fetus from potentially harmful inflammatory responses during critical stages of development. Furthermore, the absence of overt clinical manifestations and evidence of active viral replication supports the hypothesis that these viral signatures do not represent acute pathogenic infections, but rather transient exposures, latent reservoirs, or persistent viral genomic fragments lacking replicative activity.

To overcome the inherent challenges of analyzing low-biomass environments, we applied a target-enriched metagenomic approach coupled with stringent filtering criteria (Eisenhofer et al., 2019). After rigorous validation procedures, viral signatures were detected in 32% of samples, representing reproducible signals distinct from background noise. The analysis of our data demonstrates the critical importance of conservative filtering strategies; for instance, reducing the RPM threshold from 1% to 0.1% artificially increased the apparent viral prevalence (HSV-1 and rotavirus), which were not validated by independent qPCR validation. These findings point to the influence of technical artifacts and index bleed-through in low-biomass datasets (Minich et al., 2018; Kennedy et al., 2023), reinforcing the requirement for orthogonal validation.

The post-filtering viral repertoire was dominated by human-associated families, including *Herpesviridae*, *Papillomaviridae*, and *Polyomaviridae*. As these are also prominent components of the vaginal virome, our data support a potential maternal origin, suggesting the existence of shared ecological niches (Campisciano et al., 2019; Happel et al., 2023). While many identified families, such as *Flaviviridae* (Zika virus), are known to cross the placental barrier (Calvet et al., 2016), our findings extend this landscape to include species not previously described in this context, such as *Pegivirus hominis* and members of *Polyomaviridae*. Given their association with mucosal environments (Woolford et al., 2007), polyomaviruses may access the amniotic compartment via ascending transmission from the lower genital tract, consistent with mechanisms proposed for papillomaviruses.

The distribution of viral families revealed a heterogeneous landscape associated with diverse fetal outcomes. Consistent with our PCR findings, *Herpesviridae* were predominantly linked to malformations, particularly cardiac defects, supporting their involvement in congenital pathology (Alidjinou et al., 2023; Kawai et al., 2025). Conversely, *Enteroviruses* (*Picornaviridae*) were detected exclusively in fetuses with malformations, including early intrauterine growth restriction, mirroring their known tropism for fetal myocardium and central nervous system (Khediri et al., 2018; Norgan et al., 2025). In contrast, the detection of *Flaviviridae* (HPgV) in a normal fetus suggests that not all gestational viral exposures culminate in adverse outcomes. We also identified low-abundance plant viruses, likely reflecting dietary or environmental circulating nucleic acids rather than true infection (Orf et al., 2023).

To assess the in utero environment and its potential association between viral presence in AF and immune system induction, we analyzed, in parallel with viral profiling, a panel of constitutive and inducible defensins. Thus, first we characterized AMP in the AF. While HBD-1 remained stable, consistent with its role as a constitutive barrier defensin independently of viral status (Kelly et al., 2013), the inducible defensins HBD-2 and HBD-3 were significantly elevated in pathological pregnancies. This suggests that subtle immune activation may accompany developmental abnormalities even in the absence of overt infection. Notably, we observed no broad AMP differences between virus-positive and virus-negative samples, suggesting that low-level viral presence is generally insufficient to trigger systemic innate activation (Tozetto-Mendoza et al., 2025). However, a subclinical infection control (V11) with high herpesvirus load exhibited a distinct signature: elevated cathelicidins and neutrophil defensins paired with low HBD-2. This pattern is consistent with a virus-specific immune modulation rather than a classical antibacterial response (Chung and Dale, 2008).

Moreover, our high-throughput proteomic profiling revealed that the intra-amniotic immune environment is primarily governed by gestational age rather than a static immunosuppressed state (Mor and Cardenas, 2010). We observed a shift from an “immune-alert” phase in the second trimester, characterized by predominant Th1/Th2-associated signaling, innate cytotoxic responses, and active leukocyte chemotaxis, toward a third-trimester profile enriched in innate inflammatory activation and tissue-remodeling pathways, likely reflecting the progressive acquisition of a proinflammatory readiness preceding parturition (Kunze et al., 2016). While fetal sex did not influence cytokine levels, clinical status did, with significant alterations in Th1/Th2-associated networks (CXCL-10, IL-12, IL-4) in fetuses with anomalies (Saito et al., 2010).

Reinforcing our AMP findings, low-level of reads did not elicit a global cytokine response across the samples, even in the presence of subclinical viral infections. Classical inflammatory mediators and key diagnostic markers routinely used in obstetric practice, including IL-6, IL-15, CXCL-8 or CXCL-10, remained largely within normal ranges, and were not consistently upregulated in viral-positive samples. These findings suggest that low-abundance viral material may coexist within the maternal-fetal interface under biologically tolerated conditions without inducing overt inflammatory activation, while also highlighting a potential diagnostic blind spot in conventional cytokine-based clinical approaches.

Specific subsets, particularly in second-trimester virus-positive samples, showed localized IL-1A activation. Conversely, some high-load samples (VR48) showed no cytokine response, highlighting a striking uncoupling between viral positive and immune signaling. The observed downregulation of immune-regulatory markers and recruitment chemokines in some virus-positive samples may point toward a state of viral-induced immune modulation or evasion designed to preserve tissue homeostasis in an immune-privileged environment. These findings provide additional evidence that the detection of viral genetic material within the amniotic compartment does not necessarily reflect active intra-amniotic infection or overt inflammatory pathology.

Instead, the combined metagenomic, PCR-based, AMP, and proteomic analyses consistently revealed a predominantly balanced immune environment characterized by subtle and individualized immunomodulatory patterns rather than coordinated pro-inflammatory activation. The AMP profiling further reinforced this interpretation. Notably, the absence of constitutive HBD-1 expression in V72, together with the lack of broad cytokine activation except for isolated IFNG and GZMB elevation, suggests that viral detection alone is insufficient to trigger a uniform innate immune response within the amniotic environment. These observations argue against a direct relationship between viral detection and pathological inflammatory activation. The localized spike of LL-37 and HNPs 1-3 in V11 and VR65 underlines a robust mucosal counter-response to clear the elevated viral burden while simultaneously attempting to mitigate potential tissue damage. Given the known immunomodulatory and tissue-protective properties of LL-37, this localized AMP upregulation may represent an attempt to simultaneously promote viral clearance while limiting collateral tissue injury within the fetal membranes and intra-amniotic compartment. Importantly, even in these samples, broad inflammatory cytokine activation remained limited, further supporting the absence of uncontrolled inflammatory pathology.

In conclusion, our findings indicate that the AF viral landscape is a dynamic mixture of maternal, placental, and environmental signals. Rather of inducing widespread immune response, these viral nucleic acids appear to contribute to mediate localized or transient immune modulation. Our study established a conceptual framework for understanding how the “silent” persistence of viruses during gestation may shape long-term neonatal immune ontogeny, underscoring the need of integrative approaches that combine viral metagenomics with functional proteomics to clarify the clinical implications of viral signatures within the intra-amniotic environment.

## Methods

### Study design and pregnant women recruitment

A total of 114 pregnant women (maternal age range: 20-47 years) were recruited from three hospitals in Seville, Spain (Virgen del Rocío University Hospital, Virgen de Valme University Hospital, and QuirónSalud Hospital) for the collection of AF samples. The study comprised two cohorts: (1) 90 women undergoing amniocentesis (13-33 weeks of gestation), which included three diamniotic twin pregnancies; and (2) 24 women undergoing elective term cesarean sections (>37 weeks), including one diamniotic twin pregnancy.

Exclusion criteria encompassed severe medical disorders, clinical infections, or the use of prescription drugs or antibiotics within two months prior to recruitment; however, the use of antiemetic medication was permitted. Severe medical conditions were defined as those potentially altering maternal immune status or the intrauterine environment, including uncontrolled endocrine/metabolic disorders, severe cardiovascular conditions, chronic infections or immunodeficiencies, and severe organ dysfunction. Notably, sample V11 consisting of AF from a diamniotic twin pregnancy complicated by premature rupture of membranes secondary to subclinical intra-amniotic infection was retained. The patient was completely asymptomatic at the time of sample collection (via amniocentesis clinically indicated for esophageal atresia) and thus met the study’s inclusion criteria.

Criteria for downstream AF analysis required the absence of macroscopic blood contamination and a minimum sample volume of 5 mL. Based on these criteria, 9 out of 117 samples were excluded. Of the final analyzed amniocentesis AF samples (n=88), 35 corresponded to normal fetuses and 53 were associated with fetal anomalies. Among the analyzed elective cesarean AF samples (n=20), 18 corresponded to normal fetuses and two to fetal anomalies (Fig. 6). Maternal and fetal clinical characteristics including maternal age, gestational age at collection, and pregnancy-related data are summarized in Table 1. Additionally, Table S1 categorizes the samples by the specific assays performed, stratified by fetal status and gestational period.

**Fig. 6.**
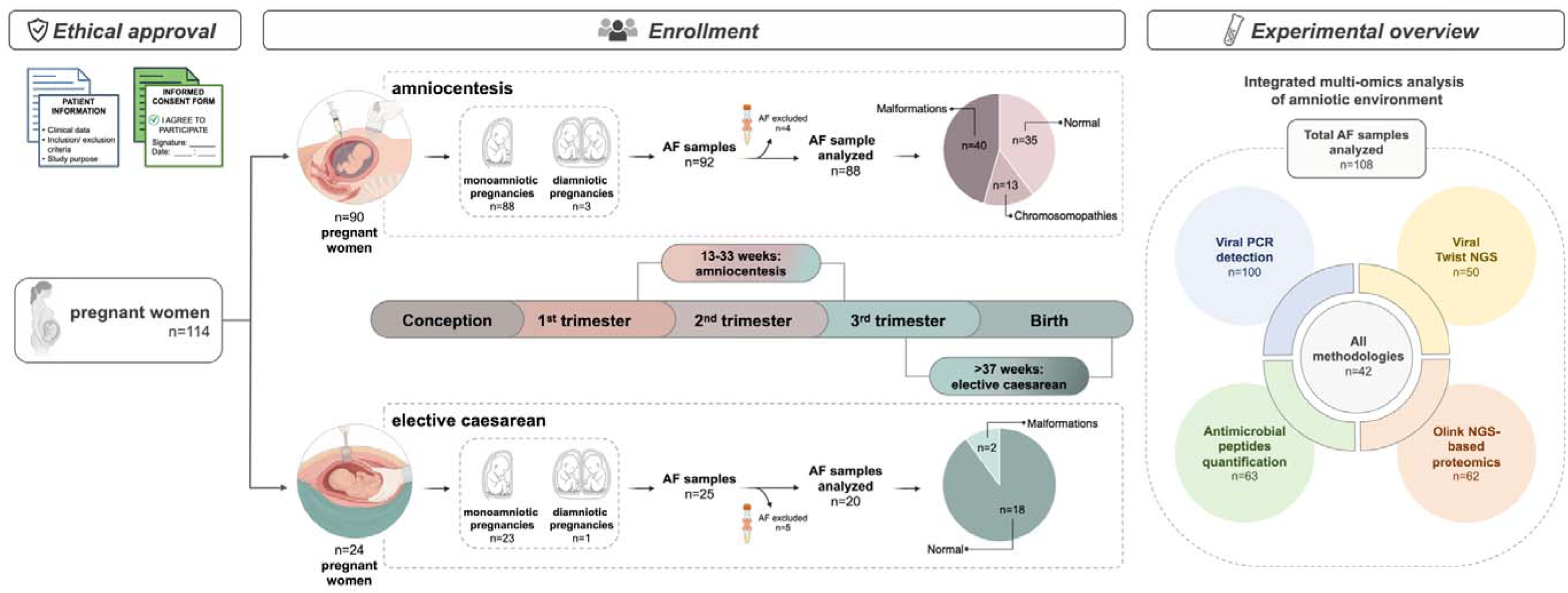
Study flowchart and experimental workflow. Overview of the sample processing and analysis pipeline for amniotic fluid samples. AF, Amniotic Fluid; NGS, Next Generation Sequencing.

**Table 1.** Baseline maternal and fetal clinical parameters of the study population. Data correspond to 108 analyzed amniotic fluid samples. W, weeks; y, years.

| Characteristics |  | Mean range or *n* (%) |  |  |
| --- | --- | --- | --- | --- |
|  |  | Amniocentesis data<br>(cohort 1) | Elective caesarean data<br>(cohort 2) | Global data |
| Maternal age (y) |  | 34.3 (20-47) | 35.8 (28-46) | 34.6 (20-47) |
| Gestational week (w) |  | 21.5 (13-33) | 38.7 (37-40) | 24.7 (13-40) |
| Fetal sex | Male | 46 (52.3%) | 12 (60%) | 58 (53.7%) |
|  | Female | 42 (47.7%) | 8 (40%) | 50 (46.3%) |
| Fetal status | Normal | 35 (39.8%) | 18 (90.0%) | 53 (49.1%) |
|  | Malformation | 40 (45.5%) | 2 (10.0%) | 42 (38.9%) |
|  | Chromosomopathy | 13 (14.7%) | 0 (0.0%) | 13 (12.0%) |

All procedures complied with the principles outlined in the Declaration of Helsinki. The study protocol was approved by the local Research Ethics Committee (REC; ID: 0452-N-22), and written informed consent was obtained from all participants prior to study inclusion.

### Amniotic fluid sample collection

AF samples (n=108) were collected aseptically by the gynecologists during amniocentesis and at term during elective caesarean section deliveries and stored at −80 °C until analysis. To minimize the potential for contamination, a rigorous aseptic protocol was followed throughout the collection and transport processes as described previously in González-Rovira et al. (2026). The collection of AF samples was conducted as follows: (1) during amniocentesis in the second trimester after abnormal findings on fetal ultrasound or biochemical marker analysis, by inserting a needle through the abdomen into the uterus to remove a 5-10 mL AF sample and, (2) during elective (planned, with no ongoing labor) caesarean delivery, at the time of caesarean section by aspirating through intact amniotic membranes using a sterile probe and 20-mL syringe.

### Viral nucleic acid extraction and quantification

For targeted detection of the *Herpesviridae* family, viral nucleic acids were extracted from 100 AF samples using the QIAamp UltraSens Virus Kit (Qiagen, Hilden, Germany; Cat. No. 3722145) according to the manufacturer’s protocol. Nucleic acids were eluted in 60 µL of AVE buffer. To monitor for potential contamination during this initial extraction workflow, three negative extraction controls (NECs) were concurrently processed using 1 mL of sterile water and eluted under identical volumetric conditions.

Concurrently, a second independent extraction workflow was established specifically for downstream shotgun metagenomic sequencing. Total DNA and RNA were extracted from 350 µL of each of 50 AF samples using the Qiagen QIAamp RNeasy Mini Kit (Qiagen; Ref. 74106). The nucleic acids were eluted in 35 µL of elution buffer, following the manufacturer’s protocol. Five NECs were processed alongside the samples using 350 µL of sterile water. The quantification of double-stranded DNA (dsDNA) was assessed using the Invitrogen Qubit 4 Fluorometer (Invitrogen, ThermoFisher Scientific, Waltham, MA, USA), according to the manufacturer’s instructions.

### Primers and DNA amplification

Two previously described sets of universal primers (Zhongliang et al., 2010) H-GF/H-GR (5′-GACTTTGCCAGCCTNTACC-3′ and 5′-GTCCGTGTCCCCGTAGATG-3′) and HU1_F/HU1_R (5′-GTGGTGGACTTTGCCAGCGTACCC-3′ and 5′-TAAACATGGAGTCCGTGTGCCGTAGATGA-3′) were modified for this study. These primers target a highly conserved region flanking a polymorphic inner sequence within the DNA polymerase gene, enabling the broad amplification of four major human herpesviruses: HSV-1/2, CMV, and EBV.

Amplifications were performed in a 25-μL final reaction volume utilizing the iStarTaq™ DNA Polymerase HotStart PCR kit (Intron Biotechnology, South Korea). The reaction mixture contained 100-1000 ng of viral DNA template, 2.5 μL of 10× PCR buffer, 1.5 μL of 25 mM MgCl2, 0.5 μL of 10 mM dNTPs, 1 μL of each forward and reverse primer (20 pmol/μL), 0.25 μL of iStarTaq™ DNA polymerase (5 U/μL), and nuclease-free water up to the final volume.

The thermocycling protocol consisted of an initial denaturation at 94°C for 5 min, followed by 40 cycles of denaturation at 94°C for 1 min, a two-step annealing profile at 53°C and 58°C (1 min each), and extension at 72°C for 1 min. A final extension step was performed at 72°C for 10 min. The resulting PCR products were resolved by electrophoresis on 2% agarose gels stained with RedSafe™ (Intron Biotechnology) and visualized under UV illumination using a GelDoc Go Imaging System (Bio-Rad, USA).

### Viral Library Preparation

Complementary DNA (cDNA) was synthesized from extracted RNA utilizing the ProtoScript II cDNA Synthesis Kit (New England BioLabs, USA). For the primer annealing step, 5 µL of random hexanucleotides (50 ng/µL) was combined with 15 µL of the RNA template and incubated at 95°C for 5 min. First-strand synthesis was subsequently performed by adding 25 µL of ProtoScript II Reaction Mix (2×) and 5 µL of Enzyme Mix (5×) to the annealed RNA. The reaction mixture was incubated at 25°C for 5 min, followed by 42°C for 1 h, and a final enzyme inactivation step at 80°C for 5 min.

For second-strand synthesis, a 30-µL master mix comprising 8 µL of NEBNext Second Strand Synthesis Reaction Buffer (5×), 4 µL of NEBNext Second Strand Synthesis Enzyme Mix, and 18 µL of molecular-grade water was added directly to the first-strand cDNA product, yielding a total reaction volume of 80 µL. Following a 1-h incubation at 16°C, the resulting double-stranded cDNA (dscDNA) was purified using AMPure XP magnetic beads at a 1.2× volumetric ratio and eluted in 25 µL of nuclease-free water.

### Target enrichment

For library construction, 25 ng of purified cDNA was processed using the Twist Library Preparation EF 2.0 Kit and the Twist Universal Adaptor System (Plates A and D; Twist Bioscience, USA). Libraries were multiplexed in pools of up to 15 samples, combining 200 ng of each individual library. Target enrichment was conducted using the Twist Comprehensive Viral Research Panel according to the manufacturer’s protocol (Tisza, et al. 2023), with probe hybridization performed at 70 °C for 16 h. Post-capture libraries were subsequently PCR-amplified for 12 cycles and sequenced on an AVITI system (Element Biosciences, USA) to generate 2 × 150 bp paired-end reads, achieving an average sequencing depth of approximately 10 million reads per sample. Following sequencing, raw binary base call (BCL) files were demultiplexed and converted to FASTQ format based on dual-index barcodes utilizing basesfastq software.

### Rotavirus and HSV-1/2 qPCR

Rotavirus detection was assessed in three samples showing high numbers of rotavirus reads by metagenomic sequencing. qPCR targeting the NSP3 gene was performed using the TaqMan Fast Virus 1-Step Master Mix (Thermo Fisher Scientific, ref. 4444432) according to the manufacturer’s instructions. Primers and probe targeting were obtained from Integrated DNA Technologies. The sequences were as follows: forward primer JVK-F, 5′-CAG TGG TTG ATG CTC AAG ATG GA-3′; reverse primer JVK-R, 5′-TCA TTG TAA TCA TAT TGA ATA CCC A-3′; and probe JVK-P, 5′-FAM-ACA ACT GCA GCT TCA AAA GAA GWG T-BHQ-3′ (Jothikumar et al., 2009). The expected amplicon size was 106 bp. qPCR was also performed to detect HSV-1 and HSV-2 in a subset of 10 samples selected based on high, medium, and low viral read counts. Reactions were carried out using the Qiagen OneStep RT-PCR Kit (Qiagen/Westburg) with virus-specific primers targeting HSV-1 and HSV-2 genomic regions. Amplification, product detection, and validation were performed following standard protocols.

### Detection of antimicrobial peptides by ELISA

AMP concentrations in 63 AF samples were quantified using highly specific enzyme-linked immunosorbent assay (ELISA) kits. The targeted AMP included α-defensins (human neutrophil peptides, HNP 1–3), β-defensins (human beta-defensins, HBD 1–3), and cathelicidin (LL-37). Specifically, HBD-1, HBD-2, and HBD-3 were measured using kits from Aviscera Bioscience (Cat# SK00858–06; Santa Clara, CA, USA), while HNP 1–3 and LL-37 were detected using assays from Hycult Biotech (Uden, The Netherlands). The validity of these specific kits for AF analysis has been previously established (Espinoza et al., 2003; Varrey et al., 2018; Para et al., 2020; González-Rovira et al., 2026). AMP concentrations were determined by interpolation from individual standard curves generated using purified peptides. Standard curve ranges were 0.97-125 pg/mL for HBD-1, 31.25-2,000 pg/mL for HBD-2, 39.5-2,500 pg/mL for HBD-3, 156-10,000 pg/mL for HNP 1-3, and 0.1-156.25 ng/mL for LL-37. The respective analytical sensitivities of the assays were 0.3 pg/mL, 7.0 pg/mL, 10.0 pg/mL, 19.46 pg/mL, and 0.14 ng/mL.

### Proteomic analysis by Olink PEA technology

Proteomic profiling of AF samples (n=62) was conducted using the high-throughput Olink® Proximity Extension Assay (PEA) platform (COBIOMIC, Córdoba, Spain), enabling the simultaneous quantification of 32 human proteins. This technology is based on an antibody-mediated proximity extension assay, wherein pairs of oligonucleotide-labeled antibodies bind to their respective target proteins (Smith and Gerszten, 2017; Haslam et al., 2022). Dual recognition of the target molecule induces oligonucleotide hybridization and subsequent extension by a DNA polymerase, generating a unique DNA barcode proportional to the initial protein abundance. These amplified products are then quantified using next-generation sequencing (NGS), providing highly sensitive and specific multiplex detection across a broad dynamic range.

Relative protein abundance was reported as Normalized Protein eXpression (NPX) values on a log2 scale, derived through Olink’s standard internal and inter-plate normalization procedures. Rigorous quality control (QC) was performed according to the manufacturer’s guidelines. This encompassed the evaluation of ten internal controls to monitor incubation, extension, and detection efficiency, alongside the assessment of sample-specific quality metrics. Only data meeting Olink’s predefined QC thresholds were retained for downstream statistical analyses.

### Bioinformatics analysis

Viral sequencing reads were processed utilizing the EsViritu pipeline (v3.2.2) via a reference-guided assembly approach. Briefly, raw reads underwent quality filtering and adapter trimming prior to alignment against reference sequences from the Virus Pathogen Database (v2.0.2). This comprehensive database encompasses all human- and animal-infecting viruses available in GenBank as of November 2025. Viral taxonomic assignments were retained only when meeting an average read identity threshold of >80%.

Data management and statistical analyses were performed using Microsoft Excel and RStudio (version 2024.09.1+394; R Core Team, 2020). The normality of data distributions was evaluated utilizing the Shapiro-Wilk test. As the data did not conform to normality assumptions, non-parametric approaches were applied. Specifically, the Mann–Whitney U test and the Kruskal-Wallis test were utilized to assess statistical significance for two-group and multi-group comparisons, respectively. A two-sided *p*<0.05 was considered statistically significant. Adjustments for multiple comparisons were performed using the Benjamini-Hochberg false discovery rate (FDR) method.

## Reporting summary

Further information on research design is available in the Nature Portfolio Reporting Summary linked to this article.

## Data availability

The sequencing data have been deposited in the NCBI database with the accession ID PRJNA1454984. Source data are provided with this paper.

## Acknowledgments

We sincerely acknowledge the valuable contribution of the gynaecologists C. López, A. López, E. De la Hoz, I. A. Castillo, A. Barranco, J. Sánchez, P. Trillo, M. D. Sánchez, F. Blanco, P. Luque, A. Maraví and B. Buezas at QuirónSalud Hospital for their essential role in the collection of amniotic fluid samples during caesarean deliveries. We are also grateful to the midwives F. Martínez and E. Flores for their coordination and handling of samples obtained at Valme University Hospital and Virgen del Rocío University Hospital, respectively. We further thank J. Swinnen and M. Bloemen for their technical assistance in sample processing and library preparation, as well as Y. Ni for support with data processing. Finally, we express our deepest appreciation to the pregnant women who generously participated in this study. This publication has been developed within the framework of the project PID2024-157768OB-I00, funded by MICIU/AEI/10.13039/501100011033 and co-financed by ERDF/EU. MK is supported by a Research Foundation Flanders (FWO) fundamental research scholarship (number: 11P7I24N). The research received financial support from the Ministerio de Ciencia, Innovación y Universidades under the same project reference. MG-R acknowledges the support of a fellowship granted by the University of Seville, Spain (PIF, VI Plan Propio de Investigación y Transferencia II.2A).

## Author contributions

Study concept and design: MG-R, EM and MLM; data acquisition: MG-R and MLM; data analysis and interpretation: MG-R, MK and MLM; technical and essential material support: LG-D, CM-P, JS, JAG-M, and JAS-B; manuscript drafting: MG-R and MLM; critical revision of the manuscript: AR-H, DB, MK, CS, EM and JM. All the authors have read and approved of the final manuscript.

## Notes

### Competing Interest Statement

The authors have declared no competing interest.

