## Supplemental Figures and Tables for "Maternal-Fetal immune networks and viral signatures in the healthy amniotic cavity"

**Table S1 | Overview of AF samples included in each experimental methodology according to fetal status and gestational trimester.** Colored boxes indicate the methodologies performed for each sample, including viral PCR detection, Viral Twist NGS, antimicrobial peptide quantification, and Olink NGS-based proteomics. Samples are grouped according to fetal outcome (normal, malformation, or chromosomopathy) and trimester of collection (1^st^, 2^nd^ and 3^rd^ trimester).

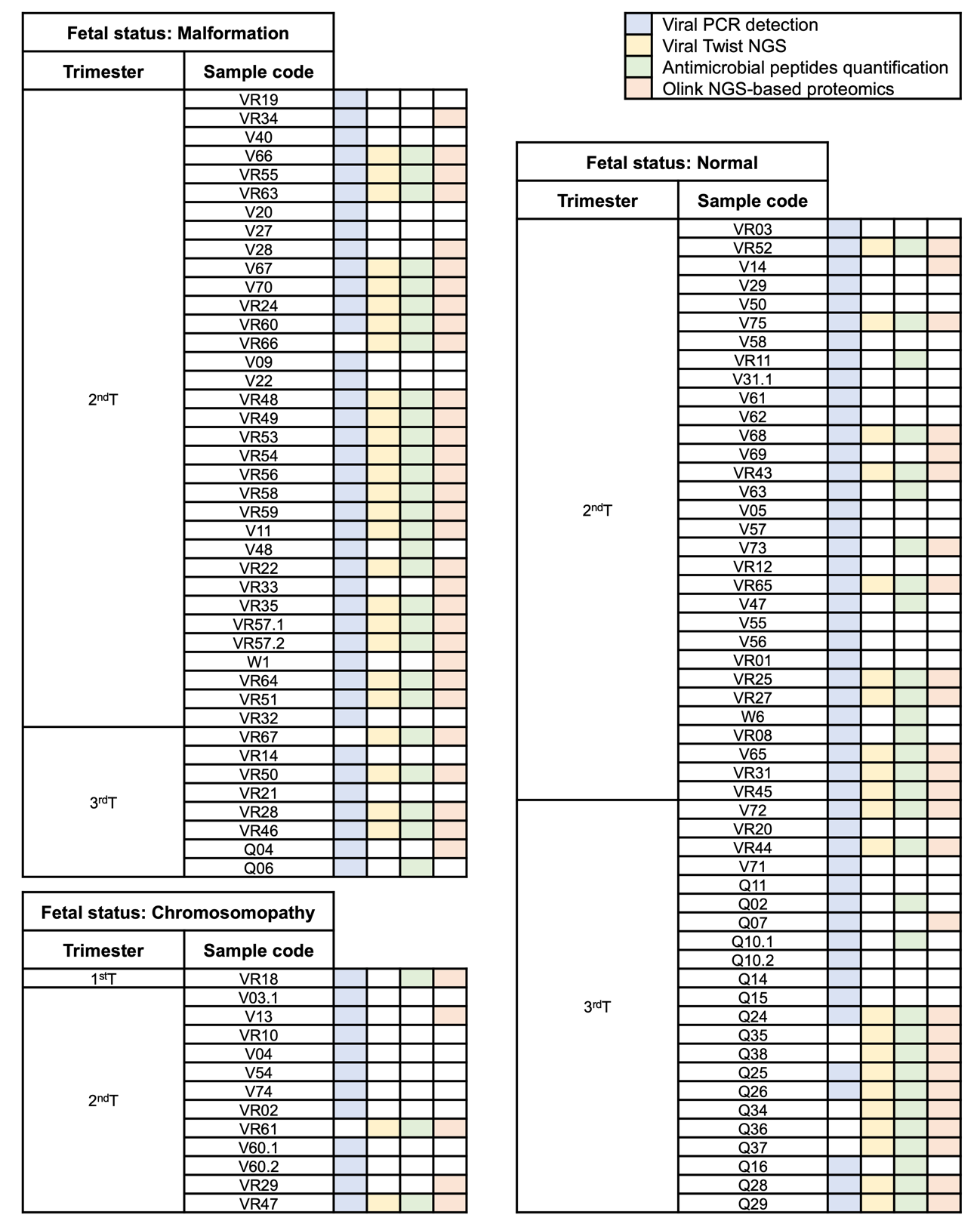

**Fig. S1 | Positive PCR amplification of *Herpesviridae* family members in amniotic fluid samples visualized by agarose gel electrophoresis.** Representative amplification products corresponding to distinct viral targets were detected at the expected fragment sizes (~500–700 bp) across amniotic fluid samples. Molecular weight marker (S) and negative control (NC) are shown. EBV, Epstein-Barr virus; HHV-6,7, *Roseolovirus humanbeta6,7*; HSV-1, *Simplexvirus humanalpha1.*

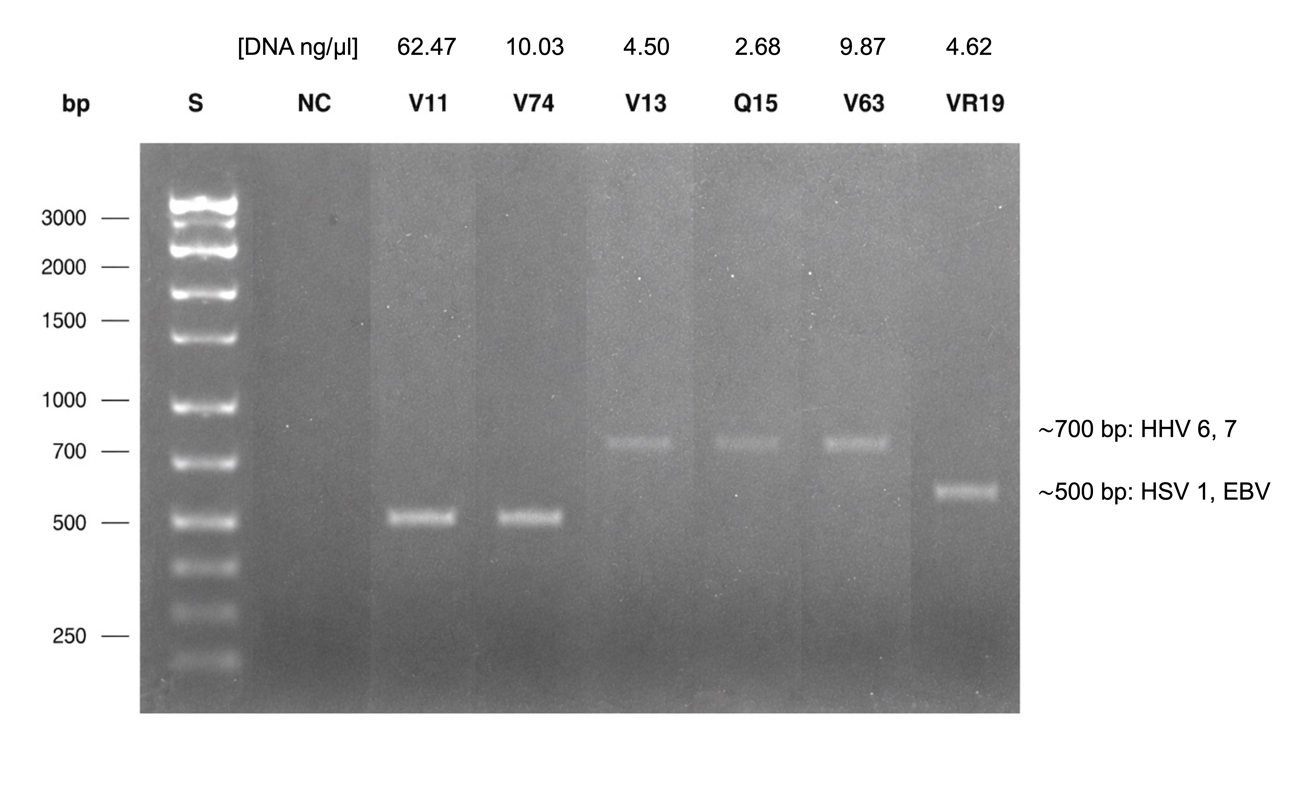

**Fig. S2 | Classification of non-human viruses detected in the intrauterine environment and associated clinical outcomes.** Plant viruses identified in 4 amniotic fluid samples, classified by genome type (RNA) and envelope status (non-enveloped), alongside their respective clinical outcomes.

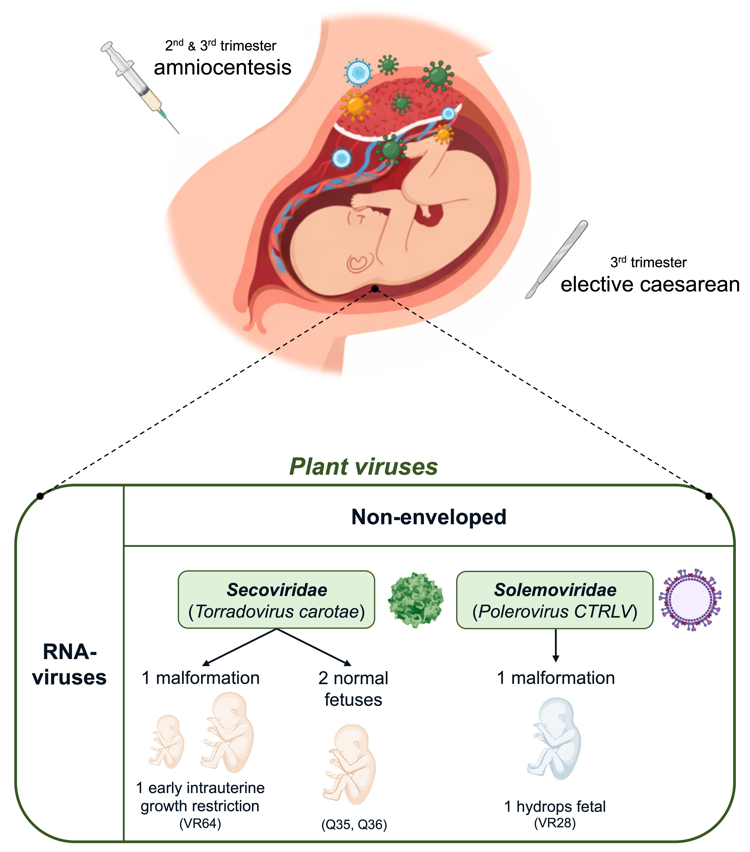

**Fig. S3 | Antimicrobial peptide concentrations in amniotic fluid according to clinical and viral features.** Concentrations of HBD-1, HBD-2, HBD-3, HNPs1-3, and LL-37 were measured in 63 AF samples. **a** Overall peptide concentration. **b-d** Peptide levels stratified by fetal sex, maternal age group, and viral detection status. **e** Peptide levels according to viral genomic type (DNA vs RNA) and envelope status (enveloped vs non-enveloped).

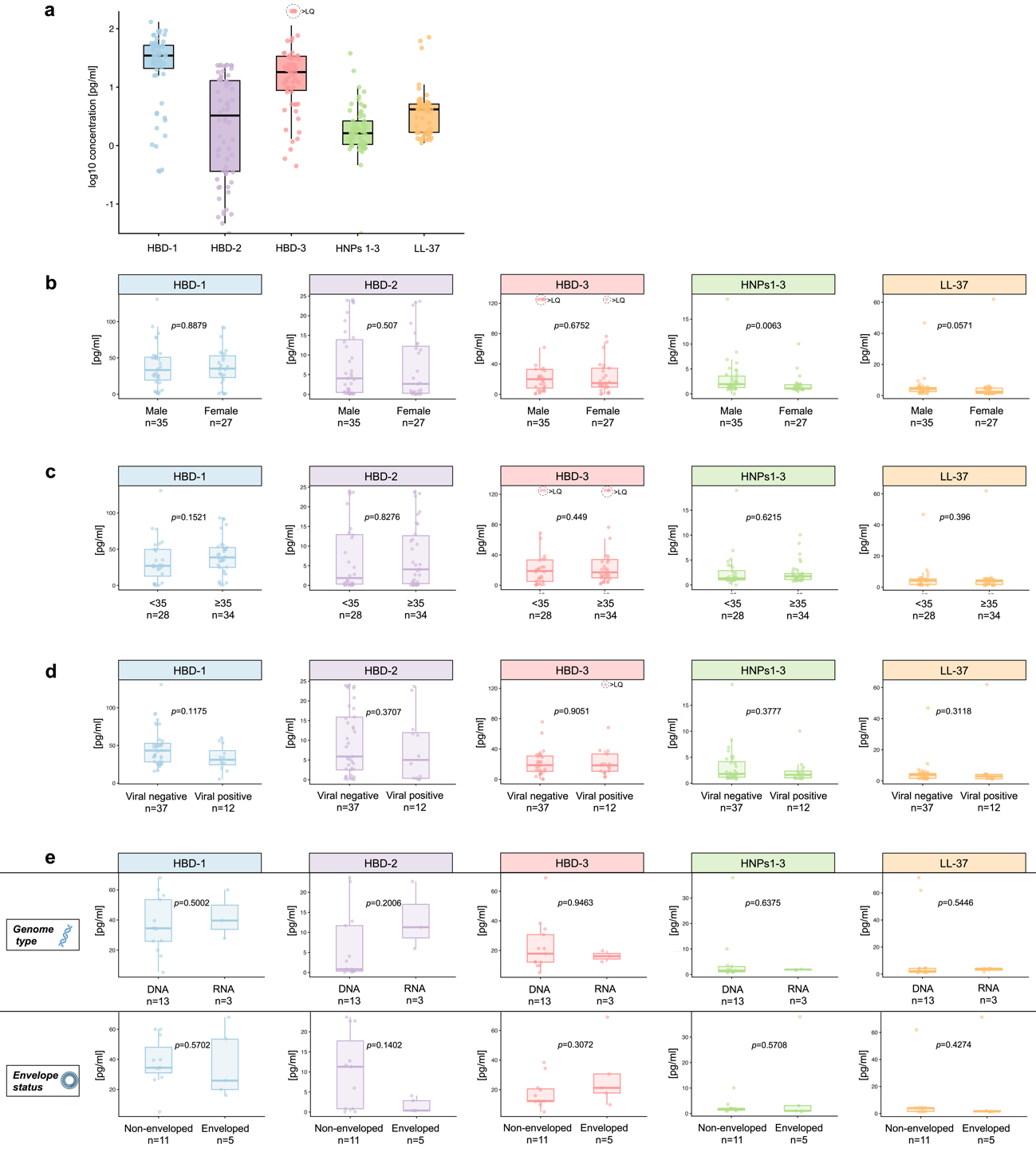

**Fig. S4 | Differential expression of immune-related proteins according to fetal status, maternal age, and viral detection.** Protein abundance was quantified using the Olink platform and is reported as normalized protein expression (NPX, log2 scale). **a** Expression levels of CXCL-10, IL-12A/IL-12B, IL-12B, and IL-4 stratified by fetal status (normal vs. abnormality fetuses). **b** IL-1B expression according to maternal age group (<35 vs. ≥35 years). **c** Expression levels of XCL-1 according to viral detection status (viral-negative vs. viral-positive samples).

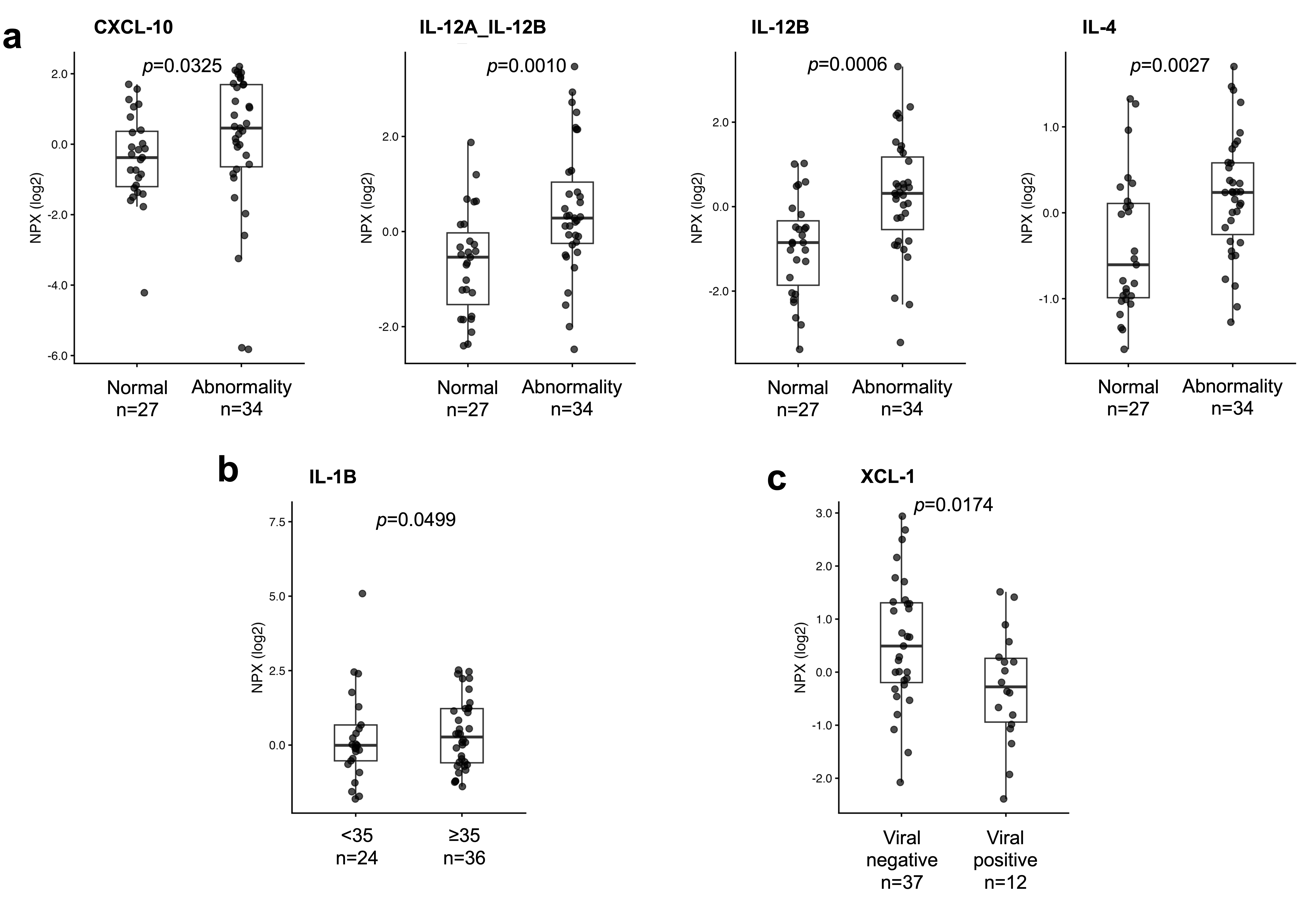
